# Does latent *Toxoplasma* infection mimic the immune profile of schizophrenia? Sex-specific cytokine and brain-marker alterations suggest partial overlap

**DOI:** 10.64898/2026.01.03.26343370

**Authors:** Jaroslav Flegr, Jana Ullmann, Filip Španiel, Jan Toman, Martin Hůla, Blanka Šebánková, Jana Petrusová, Petr Novotný, Josef Včelák, Šárka Kaňková

**Author notes:** Corresponding author: Jaroslav Flegr.

## Abstract

**Background:** Schizophrenia often features low-grade neuroinflammation. Because latent toxoplasmosis (LT) is common in this population, we tested whether LT yields a biomarker pattern resembling that reported in schizophrenia.

**Methods:** We quantified 15 cytokines and 15 blood markers of brain injury in 65 LT-positive individuals and 103 matched LT-negative controls using multiplex immunoassays. Multivariate effects of infection, age, sex, and their interaction were assessed by MANCOVA and PERMANOVA. Effects on individual biomarkers were tested by partial Kendall correlation (controlling for age and sex). Differences in the internal correlation structure were evaluated with Mantel tests on dissimilarity matrices derived from partial correlations.

**Results:** LT was associated with higher KLK6, S100B, and TDP-43 and lower MIF; several other markers showed nonsignificant but sizable trends. Cytokines showed reduced IFN-γ, IL-1β, and MCP-1 and elevated IL-13 and IL-17 in the infected group. Sex-stratified analyses suggested stronger effects on brain-injury markers in women and on cytokines in men. Correlation structure also diverged: infected individuals exhibited more negative links between brain-injury markers and cytokines, whereas controls showed predominantly positive associations (Mantel r = 0.461, p = 0.043). The LT profile overlapped with schizophrenia in elevated KLK6 and S100B and, in men, reduced GDNF, but contrasted for MIF and for the overall cytokine pattern (no consistent IL-6/TNF-α elevation).

**Conclusions:** LT entails neuroinflammatory and neuroimmune alterations that only partly recapitulate schizophrenia; the biomarker pattern and interrelationships differ, arguing against LT as the main driver of schizophrenia-related neuroinflammation.

## 1. Introduction

*Toxoplasma gondii* (Nicolle et Manceaux, 1908), a parasitic protozoan estimated to infect approximately one-third of the global population, is the causative agent of several clinically significant disease manifestations (Tenter et al. 2000, Pappas et al. 2009). The most clinically serious form is often considered to be congenital toxoplasmosis. It arises when a pregnant woman acquires the infection and transmits it to the developing fetus. Depending on the trimester in which the infection occurs, the consequences may range from miscarriage to varying degrees of fetal damage. Postnatally, infected infants may suffer from sensory and cognitive impairments, as well as severe neurological manifestations such as encephalitis, chorioretinitis, hydrocephalus, microcephaly, and intracerebral calcification (Jones et al. 2001b). Among other severe manifestations observed in specific cases is ocular toxoplasmosis caused by postnatal infection with virulent strains – particularly prevalent in regions such as South America – which may lead to blindness (Miyagaki et al. 2024). In immunocompromised individuals, such as AIDS patients or immunosuppressed organ transplant recipients, cerebral toxoplasmosis may develop after infection, which is potentially fatal. This condition requires urgent medical intervention and is otherwise often fatal (Aerts et al. 2024).

In immunocompetent individuals, acute toxoplasmosis usually progresses over the course of several weeks to a month and is often accompanied by cervical lymphadenopathy and a range of nonspecific symptoms like fever, joint pain, headache and tiredness (Jones et al. 2001a). The infection then spontaneously transitions into a chronic phase. During this phase, the parasite persists throughout the host’s lifetime as slowly replicating bradyzoites within so-called tissue cysts in various organs, including the brain. This chronic stage of infection is mostly considered asymptomatic (Markell et al. 1999, Roberts and Janovy 2000) and is commonly referred to as latent toxoplasmosis. However, the term “latent” is somewhat misleading, as infected individuals exhibit a range of more or less specific symptoms. Among the best-documented are behavioral changes, alterations in psychological profiles, and usually – but not always – impaired cognitive performance, for comprehensive review, see (Latifi and Flegr 2025). Over the past 30 years, these symptoms have primarily been investigated in the context of the manipulation hypothesis, which proposes that the parasite modifies host behavior to enhance its own transmission. In the past decade, however, it has become increasingly evident that many of the observed changes are unlikely to result from parasite-induced manipulation. Instead, they may represent side effects of deteriorated health in infected individuals (Flegr et al. 2024).

Since the mid-20th century, it has been known that *Toxoplasma*-infected individuals show increased prevalence of certain psychiatric disorders, especially schizophrenia. Initially anecdotal, these observations have since been supported by targeted epidemiological studies and, eventually, by several meta-analyses (Torrey et al. 2007, Torrey et al. 2012, Sutterland et al. 2015, Contopoulos-Ioannidis et al. 2022). Moreover, it has become clear that individuals with latent toxoplasmosis not only have an increased risk of mental health disorders but also a higher likelihood of physical health problems and exhibit numerous symptoms indicative of poorer general health. A large cross-sectional study involving 333 *Toxoplasma*-infected and 1,486 uninfected individuals found significantly worse outcomes on 28 out of 29 health-related measures in the infected group (Flegr and Escudero 2016). These individuals reported more frequent use of medications, hospitalizations, and medical complaints, with the strongest associations seen for musculoskeletal, neurological, immune, and metabolic disorders.

Similarly, an ecological study using WHO data from 88 countries found that *Toxoplasma* seroprevalence was significantly associated with disease burden in 23 of 128 disease categories and with mortality in 12 categories (Flegr et al. 2014a). For example, robust correlations were observed for cardiovascular disease, perinatal conditions, epilepsy, and certain endocrine and psychiatric disorders. In European countries, seroprevalence accounted for up to 23% of the variance in overall disease burden. In addition to these statistical findings, the study included an extensive literature review on the health effects of *Toxoplasma* infection. These and numerous other results demonstrate that latent toxoplasmosis may not be truly asymptomatic, as it has both direct and especially indirect effects on the health of infected individuals. Many of these effects may be related to changes in immune system function.

The existence of pronounced immunological changes associated with both acute and chronic *Toxoplasma* infection has been demonstrated in numerous studies. In acutely infected women, Matowicka-Karna et al. (Matowicka-Karna et al. 2009) reported elevated levels of IL-6, IL-5, and a fivefold increase in IL-10, with no significant changes in IL-12 or TNF-α – suggesting a shift toward a Th2-dominated, anti-inflammatory immune profile. Mohyuddin et al. (Mohyuddin et al. 2023) observed that in older adults, higher anti-*Toxoplasma* antibody titers were associated with increased kynurenine-to-tryptophan ratio and elevated soluble TNF receptor II, indicating chronic immune activation. In pregnant women with chronic *Toxoplasma* infection, elevated levels of IL-1β, IL-2, FGF-basic, and PDGF-BB were found (Ullmann et al., 2025). Another study in pregnant women showed increased TNF, IL-6, and IL-10 levels – but only among those with comorbid depression (Lopes et al. 2025).

Flegr and Stříž (Flegr and Stříž 2011) showed that chronically infected men had lower counts of NK cells, monocytes, and leukocytes, whereas infected women had elevated counts of these cells. Both sexes showed a reduction in B-cell numbers. Interestingly, the same study found lower *Toxoplasma* seroprevalence among men with certain immune-mediated diseases, potentially reflecting immunosuppressive effects of the parasite that mitigate immunopathology.

Also, in vitro and in vivo studies in mice have demonstrated that TNF-α, IFN-γ, and IL-12 help control parasite replication, while IL-10 may subsequently contribute to the downregulation of inflammatory mediators, including IL-2 and nitric oxide, depending on cellular context (Roberts et al. 1995, Kaňková et al. 2010, Doherty et al. 2024). Consistent with these immune dynamics, Egorov et al. (Egorov et al. 2021) reported elevated CRP and sVCAM-1 levels in infected individuals.

Altogether, the findings suggest that chronically infected individuals experience mild, chronic systemic inflammation, which may also extend into brain tissue. In support of this hypothesis, histological studies have revealed inflammatory lesions in the brain. Notably, these lesions are not co-localized with tissue cysts but are instead located near blood vessels, suggesting that the observed neuroinflammation is likely driven by immune responses rather than directly cyst-induced damage (Berenreiterova et al. 2011).

At the same time, the inflammatory response during chronic *Toxoplasma* infection appears to be tightly regulated in a way that may limit – though not necessarily prevent – immune-mediated damage to brain structure or function. Infected hosts exhibit strong pro-inflammatory responses (e.g. IFN-γ, TNF-α), but also elevated levels of some anti-inflammatory cytokines. For example, Cekanaviciute et al. (2014).demonstrated that TGF-β signaling is activated in astrocytes during toxoplasmic encephalitis and that blocking this pathway leads to greater immune cell infiltration and neuronal injury. This suggests the possible existence of an intrinsic check on inflammation within the CNS, potentially preventing excessive neuroinflammation.

It remains unclear whether this immune downregulation primarily serves the parasite – by maintaining a dormant yet transmissible bradyzoite population – or represents a host adaptation aimed to limit neuroinflammation-associated damage. One interpretation is that the parasite actively induces or exploits anti-inflammatory pathways to promote its own survival. The latent bradyzoite stage of *T. gondii* encysted in neurons is thought to be an immune-evasion strategy – a way for the parasite to persist in a dormant, transmissible form while avoiding complete immune clearance (Eberhard et al. 2025). Most likely, immune system dynamics in infected individuals result from a complex host-parasite interplay.

However, this delicate equilibrium may come at a cost. Even when overt pathology is avoided, chronic neuroinflammation and subtle immune dysregulation could affect brain function in more insidious ways. One striking example is the robust association between *Toxoplasma* infection and schizophrenia. Earlier meta-analyses reported total odds ratios as high as 2.72 (Torrey et al. 2007, Torrey et al. 2012), a considerably greater effect than that of any genetic factor identified to date. More recent studies typically report lower effect sizes (Sutterland et al. 2015, Contopoulos-Ioannidis et al. 2022). This most likely reflects the fact that, given the current emphasis on patient rights, individuals with the most severe symptoms of schizophrenia are frequently excluded from scientific studies due to their unwillingness or inability to provide informed consent (Flegr et al. 2014b). Yet, such severe cases are strongly overrepresented among individuals infected with *Toxoplasma* (Wang et al. 2006, Amminger et al. 2007, Horáček et al. 2012, Holub et al. 2013). On top of that, *Toxoplasma gondii* infection is also associated with broader structural and functional brain alterations, further implicating its role in the neurobiology and pathophysiology of schizophrenia (Horáček et al. 2012, Andreou et al. 2025).

There is a plausible mechanistic explanation for the observed association between *Toxoplasma* infection and schizophrenia. Many symptoms of schizophrenia have been linked to increased dopamine concentrations in specific brain regions. As early as 2003, it was proposed that certain behavioral effects of latent toxoplasmosis – particularly personality trait alterations – may also be related to elevated dopamine levels, and that this could represent the mechanistic link between *Toxoplasma* infection and schizophrenia (Flegr et al. 2003). This hypothesis was supported several years later by the discovery that *Toxoplasma* possesses two unique genes encoding enzymes homologous to tyrosine hydroxylase, which may affect host dopamine metabolism (Gaskell et al. 2009). A follow-up study showed high dopamine concentrations in tissue cysts and surrounding neural tissue (Prandovszky et al. 2011).

An alternative to dopamine-centered models may lie in convergent inflammatory mechanisms. Schizophrenia is associated with marked immune system abnormalities (Meyer et al. 2011, Aricioglu et al. 2016), many of which resemble those induced by *Toxoplasma gondii*. Increasing evidence suggests that innate immunity–driven inflammation represents a final common pathway in schizophrenia (Jacomb et al. 2018, Özdin and Böke 2019, Llorca-Bofí et al. 2024). *T. gondii* infection has been shown to activate the innate immune system, promoting the release of proinflammatory cytokines such as IL-1β, IL-6, TNF-α, and IFN-γ (Sher et al. 2017).

Because acute psychosis, a symptom associated with schizophrenia, is similarly characterized by immune activation, recent or reactivated toxoplasmosis may contribute to its pathophysiology.

Supporting this link, a meta-analysis of 16 studies found significantly higher odds of IgM seropositivity in acutely psychotic patients, indicating an association between recent *T. gondii* infection and acute psychosis (Monroe, Buckley, & Miller, 2015).

Any potential link between *Toxoplasma gondii* and schizophrenia should be considered within the broader context of immune-inflammatory mechanisms implicated in the disorder. Meta-analytic study of Miller et al. (2011) reported elevated levels of multiple pro- and anti-inflammatory cytokines, namely IL-6, IL-12, TNF-α, IL-1β, interferon-γ (IFN-γ), and transforming growth factor-β (TGF-β), in the blood and cerebrospinal fluid of patients with schizophrenia. Consistent with this immune activation, patients often have increased absolute and relative counts of total white blood cells, particularly monocytes (Meyer et al. 2011).

Biomarkers of astrocyte activation, such as protein S100B, provide further evidence of neuroinflammation. The presence of activated microglia in certain brain regions (notably the hippocampus) has been demonstrated using multiple techniques, including postmortem analyses and neuroimaging (PET scans) (Fineberg and Ellman 2013). Further evidence comes from flow cytometry of patients not previously treated with antipsychotic medications (i.e., drug-naïve individuals) experiencing acute psychosis, which have shown elevated CD4⁺ T cells and B cells, reduced CD8⁺ T cells, and increased plasma IL-6 (Zhuo et al. 2023). It should be emphasized, however, that these immunological findings are not uniform. A growing body of review literature points to considerable variability in immune profiles across studies, with cytokine and immune cell alterations showing dependence on clinical phase, treatment exposure, and possibly other yet-unidentified confounding variables (Miller et al. 2011, Ayyildiz et al. 2019, Reale et al. 2021).

Patients with schizophrenia also exhibit alterations in several biomarkers of neural injury, many of which are closely linked to neuroinflammatory responses or tissue repair processes. Elevated levels have been reported for Fibroblast Growth Factor 21 (FGF-21), a hepatokine that regulates glucose and lipid metabolism (Qing et al. 2015); Kallikrein-6 (KLK6), a brain-expressed serine protease involved in amyloid metabolism and upregulated during neuroinflammation; Macrophage Migration Inhibitory Factor (MIF), a pleiotropic cytokine with roles in immunoregulation and neuroprotection (Yu et al. 2022); and S100 calcium-binding protein B (S100B), an astrocyte-derived molecule widely regarded as a marker of glial activation, neural damage, and neuroinflammation (Aleksovska et al. 2014, Schümberg et al. 2016).

Conversely, patients with schizophrenia typically exhibit reduced levels of Brain-Derived Neurotrophic Factor (BDNF), a key molecule involved in neurogenesis, synaptic plasticity, and neuronal survival (Gören 2016, Liberona et al. 2024); Ciliary Neurotrophic Factor (CNTF), a neurotrophic factor with protective effects on neurons and retinal cells (Liu et al. 2020); Glial Cell Line-Derived Neurotrophic Factor (GDNF), essential for the survival and function of dopaminergic neurons (Davarinejad et al. 2025); and Ubiquitin C-Terminal Hydrolase L1 (UCHL1), a neuron-specific protein involved in removing damaged proteins (Demirel Ö et al. 2017).

The overall immune profile in schizophrenia is characterized by immune activation – manifested by elevated pro-inflammatory cytokines, activated microglia, and abnormal lymphocyte responses – and is paralleled by structural and functional brain alterations. It is important to note, however, that only 30–40% of schizophrenia patients, depending on the markers used and the patient subgroup examined, exhibit clear signs of inflammation (Murphy et al. 2021). This raises the possibility that the inflammatory subgroup may overlap substantially with individuals infected with *Toxoplasma* (Příplatová et al. 2014).

Comparing the immune alterations observed in schizophrenia with those in *Toxoplasma*-infected individuals reveals that patients with schizophrenia exhibit a range of immune system changes that significantly overlap with those seen in latent or chronic T. *gondii* infection. Both conditions are marked by a pro-inflammatory milieu (e.g., elevated IL-6, IL-1β, TNF-α), Th1-type immune activation (IFN-γ-driven), and microglia-associated neuroinflammation. While immune abnormalities in schizophrenia have been studied in great detail and documented in numerous systematic studies, focused research on the immune effects of chronic *Toxoplasma* infection in humans remains in its early stages. Furthermore, whereas schizophrenia has been linked to various molecular markers of brain injury, comparable data on corresponding brain injury biomarkers in *Toxoplasma*-infected humans are limited and fragmentary.

The aim of the present study was to test the hypothesis that previously reported differences in cytokine levels and brain injury biomarkers between individuals with and without schizophrenia might be partly explained by the higher prevalence of latent *Toxoplasma gondii* infection in patients with schizophrenia. To this end, we measured and compared the concentrations of cytokines and brain injury markers in the blood of 65 individuals chronically infected with *T. gondii* and 103 uninfected controls. We then examined whether the differences observed between infected and uninfected participants were consistent with those reported in the literature for individuals with and without schizophrenia. Finding such a concordance would support the hypothesis that some of the reported biomarker differences associated with schizophrenia could result from a higher prevalence of latent toxoplasmosis in this patient population.

## 2. Methods

### 2.1. Participants

Most participants were former students of Faculty of Science, Charles University, who had previously taken part in studies investigating the effects of *Toxoplasma gondii* infection on human behavior. We primarily invited individuals with a documented history of *Toxoplasma* seropositivity, as determined by earlier serological testing. The final sample was then supplemented with seronegative individuals to reach the target sample size, ensuring that age and gender distributions were matched between the two groups.

As a result, the proportion of infected individuals in the sample does not reflect the seroprevalence in the general population. For further details on the participants, see Section 3.1, Descriptive Statistics.

Potential participants were contacted via email and invited to the Faculty of Science at Charles University. After signing informed consent, they completed a battery of performance-based tests as part of a separate longitudinal study. A trained physician then collected venous blood samples for serological analyses.

Serum samples were obtained after centrifugation for 2 min at 3000x g at 21°C. Halt Protease and Phosphatase Inhibitor Cocktail (Thermo Fisher Scientific Inc., Waltham, MA USA) were added to the serum samples for biomarker and cytokine analyses. All serum samples were immediately stored at -20°C until analyzed. Participants received financial compensation of 500 CZK (20 EUR) for their involvement.

A total of 168 participants without acute health problems took part in the study. Serum samples from 162 participants (six samples were excluded due to test kit capacity, specifically those for whom no optimal *Toxoplasma*-positive counterpart of the same sex and similar age could be identified) were tested for brain injury biomarkers, but one participant was excluded retrospectively after showing extreme concentrations (up to 100-fold higher than average) of multiple brain damage biomarkers, suggesting a possible early-stage Alzheimer’s disease diagnosis. Serum samples from 165 participants were tested for cytokine levels (three samples were excluded due to insufficient serum volume).

The study was approved by the Institutional Review Board of the Faculty of Science, Charles University (approval number 2021/4), and conducted in accordance with applicable regulations and ethical guidelines.

### 2.2. Immunoassays

Concentrations of 15 human blood biomarkers associated with brain injury, stress, or neuroplasticity: Amyloid-β (Aβ), Brain-Derived Neurotrophic Factor (BDNF), Ciliary Neurotrophic Factor (CNTF), Fibroblast Growth Factor 21 (FGF-21), Glial Cell Line-Derived Neurotrophic Factor (GDNF), Kallikrein-6 (KLK6), Macrophage Migration Inhibitory Factor (MIF), Neural Cell Adhesion Molecule 1 (NCAM-1), Neurogranin (NRGN), S100 Calcium-Binding Protein B (S100B), TAR DNA-Binding Protein 43 (TDP-43), Total Tau (Tau, Total), Phosphorylated Tau (Tau, pT181), Ubiquitin C-Terminal Hydrolase L1 (UCHL1), and Chitinase-3-like protein 1 (YKL-40) were measured using the commercially available ProcartaPlex™ Human Neuroscience Panel, 18-plex (Cat. No. EPX180-15837-901,

ThermoFisher Scientific). This multiplex immunoassay is based on bead-based Luminex xMAP technology, which allows simultaneous quantification of multiple analytes in small-volume samples. Three markers from the original 18-plex panel were excluded from further analysis.

One (Glial fibrillary acidic protein, GFAP) was omitted due to the manufacturer’s inability to supply the necessary reagent, and two others (NF-H and NGF_beta) exhibited near-zero concentrations across almost all samples.

Similarly, concentrations of 15 human cytokines and chemokines: Granulocyte Colony-Stimulating Factor (G-CSF), Granulocyte-Macrophage Colony-Stimulating Factor (GM-CSF), Interferon-gamma (IFN-γ), Interleukin-1 β (IL-1β), Interleukin-5 (IL-5), Interleukin-6 (IL-6), Interleukin-7 (IL-7), Interleukin-8 (IL-8), Interleukin-10 (IL-10), Interleukin-12 (IL-12), Interleukin-13 (IL-13), Interleukin-17 (IL-17), Monocyte Chemoattractant Protein-1 (MCP-1), Macrophage Inflammatory Protein-1 β (MIP-1β), and Tumor Necrosis Factor-α (TNF-α) were measured using the commercially available Bio-Plex Pro Human Cytokine Panel 17-Plex (Cat. No. M5000031YV Bio-Rad). Two cytokines from this 17-plex panel (IL-2, IL-4) were excluded from analysis because their concentrations were consistently undetectable (near-zero) across almost all samples.

The presence of anti-*Toxoplasma gondii* IgG antibodies was assayed using an ELISA IgG kit (TestLine Clinical Diagnostics, Brno, Czech Republic). Examinations were carried out at the National Reference Laboratory for *Toxoplasma* infection at the National Institute of Public Health in Prague.

All assays were performed according to the manufacturers’ instructions. Detection limits, intra-assay, and inter-assay coefficients of variation for all kits are provided in the manufacturer’s documentation.

### 2.3. Statistical Analysis

All statistical analyses were performed using R version 4.4.2. Descriptive statistics (means, standard deviations, counts, and percentages) were calculated for the study cohort characteristics. Differences in age between males and females, and between infected and non-infected individuals, were assessed using independent samples t-tests. Differences in *Toxoplasma* prevalence between males and females were assessed using Pearson’s Chi-square test.

A global Multivariate Analysis of Covariance (MANCOVA) was initially performed to assess the effects of age, *Toxoplasma* infection status (infected vs. non-infected), sex, and infection-sex interaction on the concentrations of blood biomarkers of brain damage and cytokines. This analysis was conducted on a subset of 146 individuals for whom complete data across all specified biomarkers and cytokines were available. Parametric MANCOVA and its assumptions (e.g., multivariate normality, homogeneity of variance-covariance matrices) were evaluated using functions from R base, dplyr, and MVN packages.

Due to observed violations of parametric MANCOVA assumptions, a robust permutational Multivariate Analysis of Variance (PERMANOVA) was subsequently performed as a non-parametric alternative to re-evaluate the multivariate effects of age (AGE), *Toxoplasma* infection status (TOXO), sex (SEX), and the interaction between *Toxoplasma* infection and sex (TOXO:SEX) on the concentrations of the 30 blood biomarkers of brain damage and cytokines. This analysis was conducted on the same 146-participant subset as above. Prior to performing the PERMANOVA, all biomarker and cytokine concentrations were rank-transformed (using base R rank() function with ties.method = ’average’) to enhance robustness against non-normal distributions and outliers. Subsequently, Euclidean distances were calculated on these rank-transformed data, generating a dissimilarity matrix that captured the overall multivariate differences between individuals. The PERMANOVA was carried out using the adonis2 function from the vegan package. The model specified was distance_matrix_rank ∼ age + SEX + TOXO:SEX + TOXO. Statistical significance was assessed using 9999 permutations, with effects tested by sequential (Type I) sums of squares (by = "terms"), meaning the contribution of each term was evaluated after accounting for the variance explained by preceding terms in the model formula.

Given the violation of parametric assumptions, the influence of *Toxoplasma* infection on individual blood biomarkers of brain damage and cytokines was further analyzed using non-parametric partial Kendall correlation tests, rank-based method robust to non-normality and outliers. This approach allowed us to utilize data from the full cohort of 168 individuals, controlling for the confounding effects of sex and age. These partial Kendall correlations were computed using Explorer v.1 script employing the tcltk2, ppcor, corpcor, pcaPP, and psych packages.

To assess whether the internal cross-correlation structure of 30 blood biomarkers of brain damage and cytokine concentrations differed between *Toxoplasma*-positive and *Toxoplasma*-negative individuals, we constructed two dissimilarity matrices (Rtoxo: *Toxoplasma*-infected) and Rctrl: *Toxoplasma*-free) by calculating 1-partial Spearman’s correlation for all pairwise biomarker combinations within each respective group. The observed Mantel correlation coefficient (r), assessing the similarity of the partial correlation-derived dissimilarity matrices (Rtoxo and Rctrl) between groups, was then calculated. To test the hypothesis that these matrices differed more than expected by random chance, we performed a permutation test. The TOXO status was randomly permuted 1000 times, generating a null distribution of Mantel r values (ri). The one-tailed p-value was determined as the proportion of permuted r_i_ values less than or equal to the observed r: p = (sum(r_i_ ≤ robserved) + 1)/(1000 + 1).

All date are available at figshare 10.6084/m9.figshare.30408286.

## 3. Results

### 3.1 Descriptive statistics

The study cohort comprised 168 individuals (97 females and 71 males). The mean age of males (31.01±8.08 years, mean ± S.D.) did not significantly differ from that of females (31.41±7.74 years), as indicated by a t-test (t_166=_0.33, p=0.740). Among males, 24 (33.80%) were *Toxoplasma*-infected, while among females, 41 (42.27%) were infected. However, this difference in prevalence was not statistically significant (Pearson Chi-square: 1.238, df = 1, p = 0.266). Age distributions did not differ significantly between infected and uninfected individuals (31.35±9.65 years vs. 31.18±6.55 years, respectively; t_166_=0.136, p = 0.892).

Data on the concentration of blood biomarkers of brain damage were available for 161 individuals, and cytokine concentration data were available for 165 individuals. Table 1 presents the mean concentrations, stratified by infection status and sex, along with the correlations of these concentrations with age.

**Table 1:** Concentration of blood biomarkers of brain damage and cytokines by infection and their association with sex and age.

|  | Non-infected |  | Infected |  | Women |  | Men |  | Sex | Age |
| --- | --- | --- | --- | --- | --- | --- | --- | --- | --- | --- |
| Variable | Mean | S.D. | Mean | S.D. | Mean | S.D. | Mean | S.D. | Tau | Tau |
| Amyloid $\beta$ | 1.1 | 1.51 | 1.43 | 2.38 | 1.23 | 1.77 | 1.23 | 2.06 | -0.029 | -0.088 |
| BDNF | 669.23 | 531.62 | 608.09 | 453.62 | 678.32 | 523.67 | 603.26 | 473.76 | -0.075 | **0.122** |
| CNTF | 6.19 | 23.35 | 8.78 | 23.26 | 8.8 | 26.18 | 5.09 | 18.82 | **-0.105** | 0.104 |
| FGF-21 | 6.35 | 23.71 | 11.72 | 32.58 | 10.45 | 30.29 | 5.77 | 23.31 | **-0.155** | 0.037 |
| GDNF | 25.5 | 103.55 | 45.81 | 135.16 | 39.89 | 132.84 | 24.79 | 91.99 | -0.066 | **0.14** |
| Kallikrein-6 | 678.28 | 251.42 | 771.24 | 301.76 | 668.77 | 260.45 | 772.99 | 283.46 | **0.18** | **-0.122** |
| MIF | 89.73 | 43.84 | 78.18 | 38.81 | 84.24 | 38.29 | 86.64 | 47.11 | 0.024 | **0.213** |
| NCAM-1 | 40174.4 | 15995.26 | 36023.78 | 15110.23 | 36016.4 | 16087.04 | 41903.53 | 14741.63 | **0.139** | -0.02 |
| Neurogranin | 412.86 | 551.04 | 489.46 | 575.85 | 339.93 | 435.38 | 575.52 | 669.77 | **0.157** | -0.043 |
| S100B | 0.64 | 1.63 | 0.93 | 1.51 | 0.6 | 1.19 | 0.95 | 1.98 | 0.036 | **-0.15** |
| TDP-43 | 149.25 | 504.88 | 301.17 | 749.92 | 247 | 679.93 | 156.74 | 514.09 | -0.087 | 0.045 |
| Tau (Total) | 6.05 | 4.67 | 6.5 | 6.17 | 5.65 | 4.44 | 6.97 | 6.17 | 0.083 | 0.034 |
| Tau (pT181) | 0.09 | 0.47 | 0.26 | 0.98 | 0.13 | 0.58 | 0.19 | 0.85 | 0.007 | 0.019 |
| UCHL1 | 53.74 | 347.14 | 35.16 | 219.43 | 82.42 | 401.26 | 0 | 0 | **-0.156** | 0.032 |
| YKL.40 | 6320.31 | 3512.14 | 5525.86 | 2656.33 | 6350.13 | 3045.78 | 5577.89 | 3415.27 | **-0.131** | **0.154** |
| G-CSF | 1.59 | 8.2 | 0.49 | 3.88 | 1.09 | 6.37 | 1.31 | 7.65 | 0.001 | **0.121** |
| GM-CSF | 0.95 | 7.37 | 0.07 | 0.45 | 0.07 | 0.47 | 1.4 | 9.06 | 0.042 | -0.057 |
| IFN $\gamma$ | 10.14 | 7.16 | 13.11 | 38.11 | 12.46 | 30.59 | 9.54 | 7.76 | -0.068 | **0.12** |
| IL-1 $\beta$ | 0.22 | 0.5 | 0.09 | 0.27 | 0.15 | 0.37 | 0.2 | 0.51 | 0.065 | 0.087 |
| IL-5 | 21.69 | 124.57 | 5.88 | 12.26 | 8.27 | 15.15 | 26.42 | 152.9 | **0.166** | 0.103 |
| IL-6 | 3.12 | 22.46 | 0.63 | 2.15 | 0.73 | 3.55 | 4.25 | 27.39 | -0.096 | 0.008 |
| IL-7 | 2.81 | 6.3 | 4.46 | 12.63 | 3.82 | 10.38 | 2.87 | 7.23 | 0.017 | 0.035 |
| IL-8 | 15.62 | 60.65 | 26.97 | 77.53 | 16.5 | 55.13 | 24.71 | 82.17 | -0.04 | 0.055 |
| IL-10 | 2.22 | 2 | 2.11 | 1.33 | 2.3 | 1.48 | 2.01 | 2.13 | **0.117** | **0.21** |
| IL-12 | 11.63 | 25.7 | 8.4 | 9.69 | 9.4 | 9.34 | 11.87 | 31.11 | **-0.164** | **-0.12** |
| IL-13 | 1.14 | 2.23 | 1.44 | 1.57 | 1.26 | 1.53 | 1.24 | 2.55 | **-0.173** | 0.086 |
| IL-17 | 0.91 | 1.33 | 1.83 | 3.69 | 1.22 | 1.64 | 1.3 | 3.42 | **-0.106** | **-0.118** |
| MCP-1 | 27.18 | 19.15 | 20.97 | 15.69 | 22.97 | 15.26 | 27.52 | 21.41 | -0.093 | 0.022 |
| MIP-1 $\beta$ | 38.84 | 56.72 | 36.54 | 39.4 | 33.41 | 39.07 | 44.49 | 63.67 | 0.072 | **0.156** |
| TNF alfa | 33.66 | 107.14 | 23.34 | 21.27 | 35.01 | 108.24 | 22.32 | 32.63 | 0.078 | 0.054 |

*All concentrations are in pg/mL. The last two columns present the partial Kendall Tau* *correlation coefficients, controlled for either age or sex, between the biomolecule concentrations* *and sex (coded 0 for women, 1 for men) or age, respectively. A positive Tau value indicates that* *men or older individuals generally exhibit higher concentrations of the biomolecule compared to* *women or younger individuals. Significant Tau values are bolded.*

### 3.2 Effects of Toxoplasma infection on concentrations of blood biomarkers of brain damage and cytokines

A global Multivariate Analysis of Covariance (MANCOVA) was performed on a subset of 146 individuals for whom data on all blood biomarkers of brain damage and all cytokines were available (Table 2).

**Table 2:**
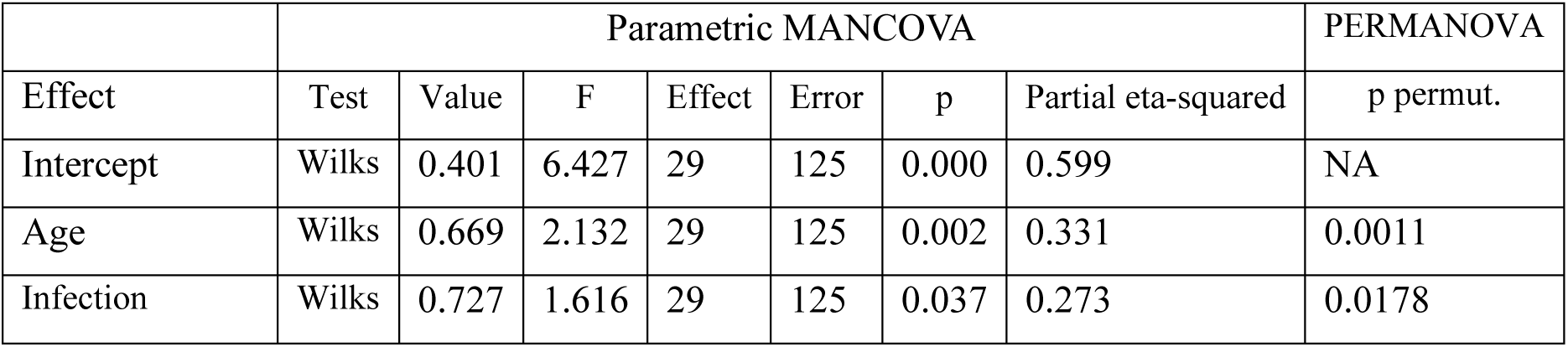
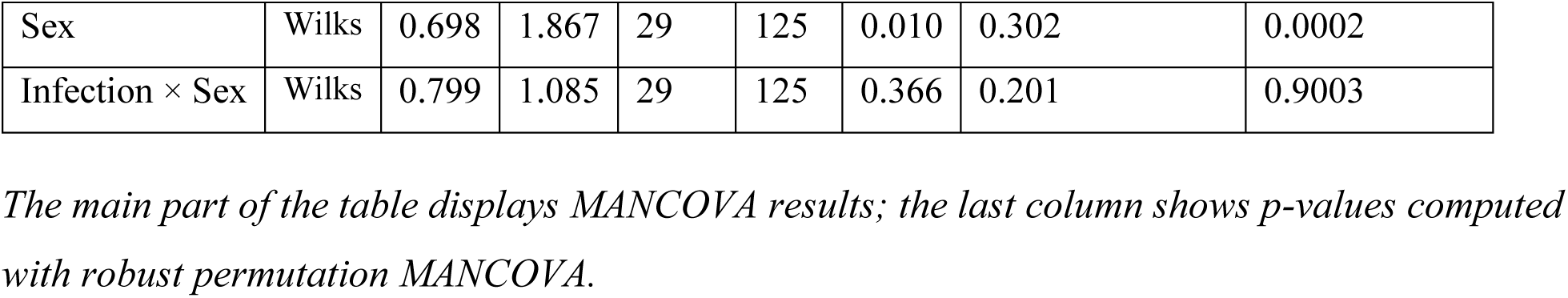
Effects of age, *Toxoplasma* infection, and sex on blood biomarkers of brain damage and cytokines.

|  | Parametric MANCOVA |  |  |  |  |  |  | PERMANOVA |
| --- | --- | --- | --- | --- | --- | --- | --- | --- |
| Effect | Test | Value | F | Effect | Error | p | Partial eta-squared | p permut. |
| Intercept | Wilks | 0.401 | 6.427 | 29 | 125 | 0.000 | 0.599 | NA |
| Age | Wilks | 0.669 | 2.132 | 29 | 125 | 0.002 | 0.331 | 0.0011 |
| Infection | Wilks | 0.727 | 1.616 | 29 | 125 | 0.037 | 0.273 | 0.0178 |
| Sex | Wilks | 0.698 | 1.867 | 29 | 125 | 0.010 | 0.302 | 0.0002 |
| Infection × Sex | Wilks | 0.799 | 1.085 | 29 | 125 | 0.366 | 0.201 | 0.9003 |

The main part of the table displays MANCOVA results; the last column shows *p*-values computed with robust permutation MANCOVA.

While the initial parametric MANCOVA indicated significant effects of Age, Infection, and Sex, but not Infection × Sex interaction, it was noted that the data violated most assumptions for the MANCOVA test. Therefore, a non-parametric alternative – permutational multivariate analysis of variance (PERMANOVA) – was used to assess the same multivariate effects. PERMANOVA also revealed significant effects for *Toxoplasma* infection (Table 2, last column). Therefore, in the subsequent phase of the study, the influence of infection on individual blood biomarkers of brain damage and cytokines was analyzed separately using non-parametric partial Kendall correlation tests, controlling for the effects of sex and age. This approach allowed us to revert to the original cohort of 168 individuals. Tables 3 and 4 present the results of these analyses for the entire cohort, as well as separately for males and females. Of the 15 blood markers of brain damage studied, the concentrations of four demonstrated significant correlations with *Toxoplasma* infection even after correction for multiple comparisons: Kallikrein-6, S100B, and TDP-43 correlated positively, while MIF correlated negatively. Several other markers exhibited non-significant but sizable trends: positive for Tau (pT181); negative for GDNF, NCAM-1, and YKL.40. The effects of *Toxoplasma* infection (as indicated by partial Kendall Tau coefficients) were predominantly stronger in women than in men (e.g., Kallikrein-6: 0.217 vs. 0.070; MIF: - 0.152 vs. -0.064; Neurogranin: 0.178 vs. -0.063; S100B: 0.211 vs. 0.070). An exception was GDNF, where the effect of *Toxoplasma* infection was not evident in women, but a significant decrease was observed in men (Tau = -0.191). The concentration of Amyloid β, a peptide associated with Alzheimer’s pathology, showed no significant association with *Toxoplasma* infection overall. However, it showed a negative trend in men (Tau = -0.096) and a positive trend (Tau = 0.078) in women.

**Table 3:**
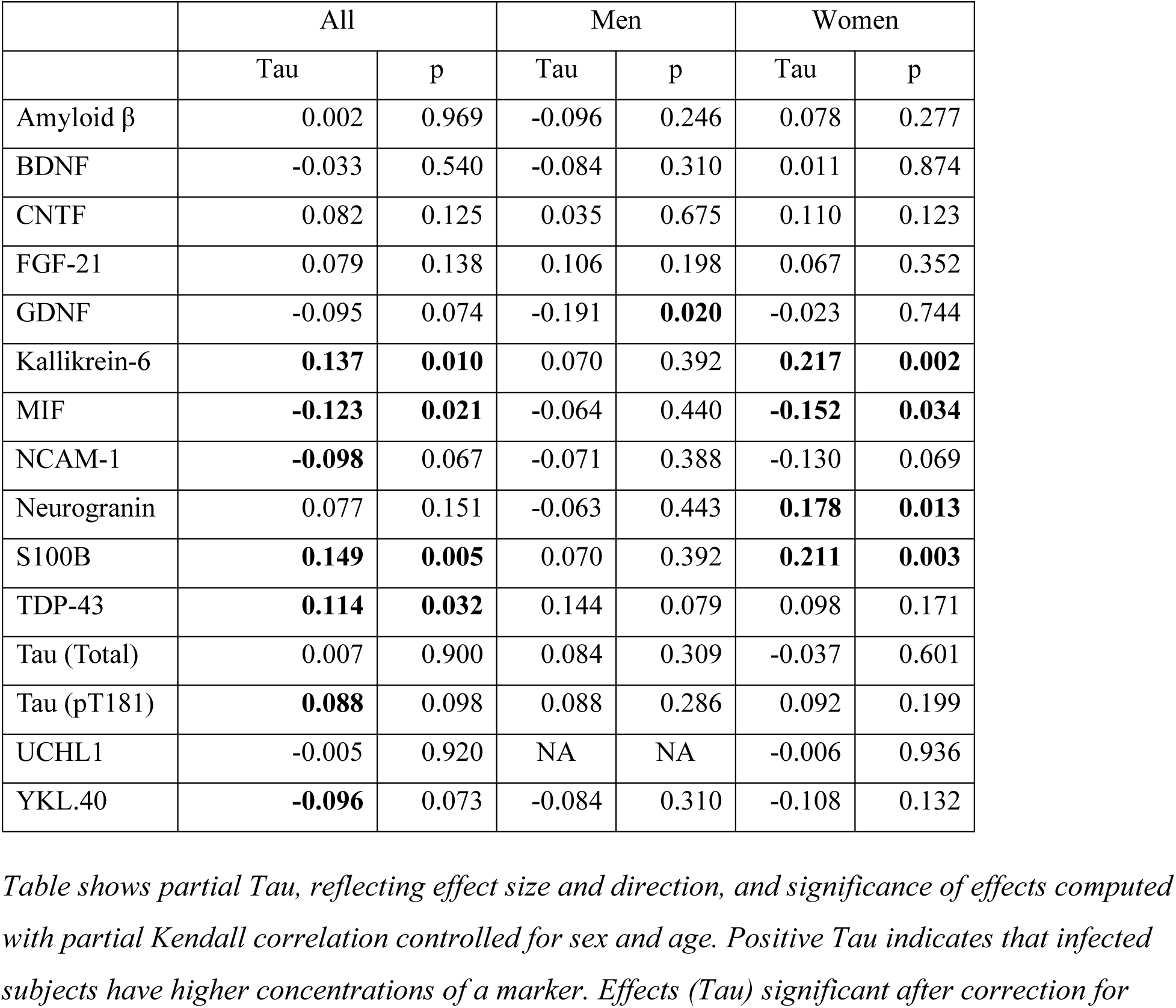
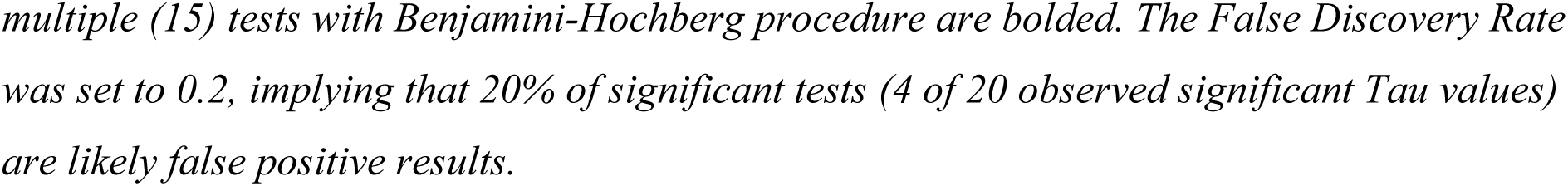
Effects of *Toxoplasma* infection on concentration of blood biomarkers of brain damage.

**Table 4:** Effects of *Toxoplasma* infection on concentration of cytokines.

|  | All |  | Men |  | Women |  |
| --- | --- | --- | --- | --- | --- | --- |
|  | Tau | p | Tau | p | Tau | p |
| G-CSF | -0.059 | 0.263 | -0.110 | 0.187 | -0.025 | 0.715 |
| GM-CSF | -0.002 | 0.964 | **-0.163** | 0.051 | 0.123 | 0.075 |
| IFN $\gamma$ | **-0.111** | **0.035** | -0.149 | 0.074 | -0.086 | 0.216 |
| IL-1 $\beta$ | **-0.114** | **0.030** | -0.056 | 0.502 | -0.164 | **0.018** |
| IL-5 | -0.077 | 0.147 | -0.063 | 0.449 | -0.078 | 0.260 |
| IL-6 | 0.074 | 0.159 | 0.027 | 0.745 | 0.110 | 0.112 |
| IL-7 | -0.013 | 0.803 | -0.027 | 0.747 | -0.004 | 0.957 |
| IL-8 | 0.000 | 1.000 | 0.027 | 0.748 | -0.014 | 0.836 |
| IL-10 | 0.016 | 0.757 | -0.031 | 0.709 | 0.057 | 0.409 |
| IL-12 | -0.020 | 0.700 | -0.027 | 0.750 | -0.027 | 0.692 |
| IL-13 | **0.153** | **0.004** | **0.200** | **0.017** | 0.130 | 0.060 |
| IL-17 | **0.132** | **0.012** | **0.178** | **0.033** | 0.106 | 0.124 |
| MCP-1 | **-0.127** | **0.016** | **-0.228** | **0.006** | -0.070 | 0.311 |
| MIP-1 $\beta$ | 0.040 | 0.450 | 0.058 | 0.491 | 0.023 | 0.743 |
| TNF alfa | 0.022 | 0.681 | -0.044 | 0.596 | 0.053 | 0.441 |

Table shows partial Tau, reflecting effect size and direction, and significance of effects computed with partial Kendall correlation controlled for sex and age. Positive Tau indicates that infected subjects have higher concentrations of a marker. Effects (Tau) significant after correction for multiple (15) tests with Benjamini-Hochberg procedure are bolded. The False Discovery Rate was set to 0.2, implying that 20% of significant tests (3-4 of 18 significant Tau values) are likely false positive results.

Table 4 illustrates the effects of *Toxoplasma* infection on cytokine concentrations. Again, it was found that 5 out of the 15 studied cytokines correlated with *Toxoplasma* infection: IFN gamma, IL-1 β, and MCP-1 correlated negatively, while IL-13 and IL-17 correlated positively. For cytokines, the effects and trends were generally stronger in men than in women, with the exception of IL-1 β, where the negative effect was approximately three times stronger in women (Tau = -0.164) than in men (Tau = -0.056, non-significant). Notably, GM-CSF concentration correlated negatively with *Toxoplasma* infection in men (Tau = -0.163) but positively in women (Tau = 0.123).

### 3.3 Data exploration

#### 3.3.1 Cross-correlation between concentration of markers and cytokines

In the next part of the study, we analyzed the correlation between the concentrations of individual biomolecules (Fig. 1). Generally, blood biomarkers of brain damage exhibited positive correlations with each other, with correlation strengths typically being high. Out of 105 pairwise comparisons, 57 showed significant positive correlations. Conversely, all 13 negative correlations were non-significant (p>0.077). Similarly, positive and strong correlations were observed among the concentrations of individual cytokines. Of 105 cytokine pairs, 89 correlated positively, with 70 being significant, and only 16 correlated negatively, with only three negative correlations (IL-8:IL-10, IL-8:IL-13, and IL-8:IL-17) reaching statistical significance.

**Fig. 1.**
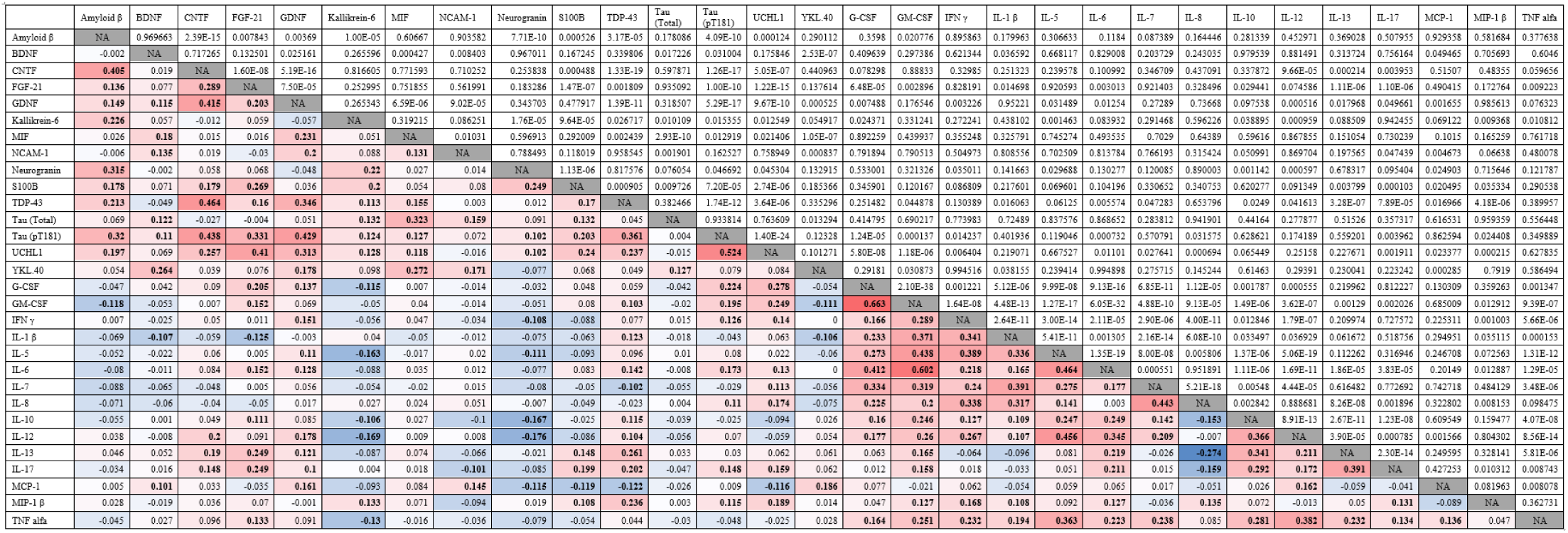
Matrix of pairwise correlations between individual blood biomarkers of brain damage and cytokines. Cells below the diagonal report Kendall’s τ (effect size and direction); cells above the diagonal show the corresponding p-values from the partial Kendall’s τ test adjusted for age and sex. Color intensity encodes |τ|: red indicates positive associations and blue negative ones (deeper colors = stronger effects). Significant τ values are bolded. No correction for multiple comparisons was applied in this exploratory analysis.

Among the 225 possible blood biomarker-cytokine pairs, 48 showed significant positive correlations and 20 exhibited significant negative correlations; only MIF and Total Tau showed no statistically significant correlations with any of the cytokines tested. Notably, similar correlation matrices calculated separately for *Toxoplasma*-infected and non-infected participants revealed substantial structural differences. For 66 infected individuals, among the 225 blood marker of brain damage-cytokine correlations, 8 significant positive correlations and 31 significant negative correlations were observed (with a similar number approaching significance). In contrast, among 108 non-infected individuals, there were 63 significant positive correlations and only 6 negative correlations (Fig. 2).

**Fig. 2.**
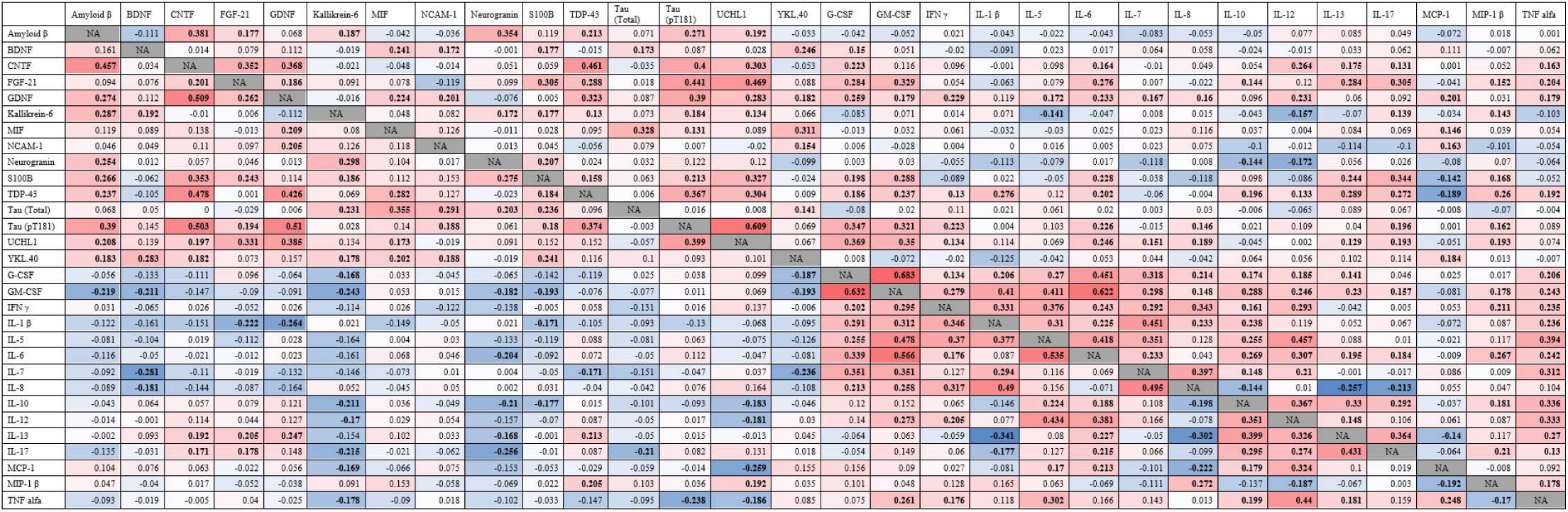
Matrices of pairwise correlations between individual blood biomarkers of brain damage and cytokines for *Toxoplasma*-infected and non-infected subjects. *Cells below the diagonal display Kendall’s Tau coefficient for the 66 Toxoplasma-infected subjects, whereas cells above the diagonal present these values for the 108 non-infected subjects. For additional information regarding the interpretation of these values, please refer to the legend of* Fig. 1.

To investigate whether the pattern of pairwise correlations among the studied biomolecules differed between *Toxoplasma*-positive and *Toxoplasma*-negative individuals, a Mantel test, a permutation-based significance test, was performed by comparing the two corresponding dissimilarity matrices. The observed Mantel correlation coefficient (r) between the *Toxoplasma*-positive and *Toxoplasma*-negative cross-correlation matrices was 0.461. A permutation test, where individuals were randomly assigned to either group 1000 times, revealed that only 0.043 (or 4.3%) of the randomly generated Mantel r values were lower than the observed r of 0.461.

This indicates a statistically significant difference in the cross-correlation matrix structures between *Toxoplasma*-positive and *Toxoplasma*-negative individuals (p = 0.043). The relatively low observed Mantel r value of 0.461, compared to the distribution of r values obtained from permuted datasets, suggests that the internal relational patterns of the biomolecules are indeed significantly less similar between the actual *Toxoplasma*-defined groups than would be expected by chance.

## 4. Discussion

This study investigated the association of *Toxoplasma* infection with blood levels of 30 analytes—15 cytokines and 15 brain-injury markers—in a cohort of 168 individuals. A parametric MANCOVA (controlling for age and sex) indicated a significant overall effect of infection; because several model assumptions were violated, we confirmed this result with a non-parametric ANCOVA (PERMANOVA). To identify which molecules contributed to the multivariate signal, we then ran post hoc partial Kendall correlations (τ), again adjusting for age and sex. Key findings include specific significant associations between *Toxoplasma* infection, multiple blood biomarkers of brain damage and a subset of cytokines. Furthermore, notable differences were observed in the patterns of correlation matrices for blood biomarkers of brain damage and cytokines when comparing *Toxoplasma*-infected and non-infected participants.

While in 103 *Toxoplasma*-free subjects, the concentration of the majority of blood biomarkers of brain damage (63 of 69) correlated positively with cytokine levels across all possible combinations of biomarkers and cytokines, among 65 *Toxoplasma*-infected individuals, the majority (31 of 39) of significant correlations were negative (see tab. 6).

Our results corroborate accumulating evidence that latent *Toxoplasma* infection is not truly asymptomatic, but is associated with chronic low-grade neuroinflammation and disruption of the neuro-immune system. The significant increase in KLK6, S100B, and TDP-43 in infected individuals may reflect low-grade cellular stress and subclinical damage within the central nervous system (CNS). S100B is a well-established marker of astrocyte activation or injury (Langeh and Singh 2021), and its elevation is consistent with the known neurotropism of *T. gondii*, which is unable to establish persistent cysts within glial cells, leading to reactive astrocytosis (Everett and Elsheikha 2025). However, under certain conditions, S100B also exhibits neuroprotective activity – for example, by reducing TNF-α production (Langeh and Singh 2021, Everett and Elsheikha 2025). The increase in KLK6, a serine protease implicated in neuroinflammatory processes and the degradation of CNS components (Patra et al. 2018, Mella et al. 2020), further supports this interpretation. The rise in TDP-43, another protein linked to neurodegenerative diseases like Amyotrophic lateral sclerosis (ALS), Frontotemporal lobar degeneration (FTLD) and Alzheimer’s disease (AD), may also signify a subclinical stress response within neurons. The significant decrease in Macrophage Migration Inhibitory Factor (MIF) is intriguing. As a potent pro-inflammatory cytokine, MIF is critical for host resistance against *T. gondii* (Flores et al. 2008, Terrazas et al. 2010). It is possible that its suppression in chronically infected individuals could reflect an evolved immune evasion strategy by the parasite, aimed at dampening an effective host response and ensuring parasites’ long-term survival. Increased, rather than decreased level of MIF was observed in schizophrenia patients (Yu et al. 2022). However, Yu et al.’s study was performed on first episode schizophrenia patients only.

Generally, the observed cytokine profile in infected individuals is paradoxical and is consistent with the hypothesis of parasite-induced modulation of host immunity. The cornerstone of protective immunity against *T. gondii* is a robust Th1 response, driven primarily by IFN-γ and IL-12 (Sturge and Yarovinsky 2014, Ihara and Yamamoto 2024). Our finding of significantly lower levels of IFN-γ, along with the pro-inflammatory cytokine IL-1β and the monocyte-recruiting chemokine MCP-1, in chronically infected subjects is counterintuitive from a host-defense perspective. However, it is in line with the known ability of *T. gondii* to actively suppress IFN-γ signaling pathways to evade clearance and establish lifelong latency (Kim et al. 2007, Rosowski et al. 2014). This suggests that the parasite establishes a state of modulated, “cold“ inflammation, sufficient to prevent uncontrolled propagation but not to achieve sterile immunity. This is further corroborated by the concurrent increase in IL-13 and IL-17. Elevated IL-13 indicates a shift toward a Th2-type response, which is known to antagonize the protective Th1 immunity (Wynn 2015). The increase in IL-17, a key cytokine of the Th17 lineage, may play a dual role: while it can be protective during acute infection, its sustained elevation in the chronic phase may promote persistent low-grade neuroinflammation and compromise the integrity of the blood-brain barrier (Cua and Tato 2010).

Perhaps the most notable finding of this study is the reversal of the correlation patterns between brain damage biomarkers and cytokines in infected versus non-infected individuals. In the uninfected cohort, the network behaves as expected: markers of cellular stress are positively correlated with inflammatory cytokines, reflecting a coordinated physiological response to maintain homeostasis. In the *Toxoplasma*-infected cohort, this network appears markedly altered, dominated by negative correlations. This suggests that latent *Toxoplasma* infection does not merely alter the levels of specific molecules but profoundly disrupts the communication and regulatory logic of the entire neuro-immune system. Such systemic dysregulation, in which the usual relationship between damage signals and immune response is disrupted, could underlie certain long-term neurological and psychiatric effects associated with *Toxoplasma* infection.

Additionally, this “rewiring“ may represent the ultimate host manipulation by the parasite, resulting in a dysregulated immune environment that favors parasite persistence while potentially impairing host neuro-immune integrity.

We observed sex-specific differences in the associations of brain damage markers and cytokines with infection. Specifically, the association between *Toxoplasma* infection and brain injury biomarkers was stronger in women, whereas cytokine alterations were more pronounced in men. These findings are in line with the growing body of literature on sexual dimorphism in immunity to *Toxoplasma* (Roberts et al. 1995, Walker et al. 1997). Hormonal differences are known to shape immune responses, with females generally mounting stronger humoral and males stronger cellular responses (Gay et al. 2021). Studies in mice have shown that males can produce IFN-γ more rapidly, potentially leading to better initial control of the parasite but different long-term inflammatory sequelae (Walker et al. 1997). The pronounced reduction of GDNF in infected men is particularly notable, given its role in supporting dopaminergic neurons (Bourque and Trudeau 2000, Mätlik et al. 2022) and the reported increase in dopaminergic activity in *Toxoplasma*-infected individuals (Flegr et al. 2003, Prandovszky et al. 2011) – a phenomenon that is itself modulated by sex-specific neuroendocrine factors (Woodcock et al. 2020, Williams et al. 2021). These findings suggest that *Toxoplasma* may exploit baseline sex-specific immune and neuroendocrine differences, resulting in distinct pathophysiological pathways and potentially different neuropsychiatric outcomes in men and women.

A potentially important sex-specific observation concerns the opposing trends in amyloid β concentrations, a key marker of Alzheimer’s disease, between infected and uninfected individuals. While the differences are not statistically significant in either sex, the directionally opposing trends are noteworthy despite relatively small effect sizes (Kendall’s Tau = −0.096 for men; 0.078 for women). This potential sex-specific divergence might help reconcile the often contradictory findings regarding the association between toxoplasmosis and Alzheimer’s disease (AD) (Bayani et al. 2019, Nayeri Chegeni et al. 2019), as well as the higher prevalence of AD in women compared to men (Fiest et al. 2016, Matthews et al. 2016, Podcasy and Epperson 2016). Some studies have also demonstrated that, while infected women tend to perform worse than uninfected controls in certain cognitive tests, infected men perform better than uninfected men on certain cognitive tasks (Flegr et al. 2012, Stock et al. 2014, Wyman et al. 2017). In this context, the results of studies using a mouse model of AD (5xFAD strain) may be relevant.

Experimental infection with *Toxoplasma gondii* was associated with a marked reduction in amyloid plaque density throughout the brain, particularly in the cortex and hippocampus (Yanes et al. 2024). This was likely due to the recruitment of Ly6Chi monocytes and enhancement of phagocytosis and degradation of soluble Aβ (Möhle et al. 2016).

Comparison of our findings with previously published data is challenging, particularly in the case of brain damage markers. A recent study – whose Russian authors explicitly declared that no dataset was generated during the study – reports highly implausible statistical results (e.g., 15 out of 34 low-powered tests significant, 13 of which reportedly reached p < 0.001 after an correction for multiple comparisons). Owing to the questionable credibility of these findings, we have chosen not to cite the study directly. However, the reference is available upon request.

For cytokines, two recent studies have been published, both conducted on pregnant women. The first, originating from our institution, reported increased concentrations of cytokines such as IL-1β, IL-1ra, IL-2, FGF-basic, and PDGF-BB in infected women (Ullmann et al. 2025). The second study found elevated levels of TNF, IL-6, and IL-10, and decreased levels of IL-8 and MIF, although these changes reached statistical significance only in the subgroup of pregnant women with depression (Lopes et al. 2025). The cytokine shifts observed in depressed women largely corresponded to our current findings in terms of direction, although not always in magnitude. In contrast, the results published in our earlier study – except for the consistently elevated TNF-α – differed from our present findings and, in some cases, even contradicted them in direction. Although our previous study employed a different cytokine detection kit from the same manufacturer, both analyses were performed in the same laboratory using the same instrumentation approximately two years apart. However, the study populations differed substantially, particularly in age, and more importantly, in pregnancy status. The earlier data were collected from pregnant women. It is well established that pregnancy induces substantial shifts in immune system functioning due to the need to maintain immune tolerance toward the semi-allogeneic fetus while preserving antimicrobial defense (Guleria and Sayegh 2007, Abu-Raya et al. 2020, Li et al. 2020).

The primary aim of our study was to seek an empirical support for the hypothesis that the increased level and incidence of neuroinflammation observed in patients with schizophrenia may be related to the elevated prevalence of latent toxoplasmosis in this population. Our findings suggest that the cytokine and brain injury biomarker profiles in *Toxoplasma*-infected individuals share notable similarities with those reported in patients with schizophrenia. Notably, elevated levels of S100B, a trend toward increased FGF-21, and a pronounced trend toward decreased GDNF levels (statistically significant in men) are fully consistent with this pattern.

However, some notable discrepancies with previously published findings were also observed. In our study, *Toxoplasma*-infected individuals exhibited decreased levels of MIF, whereas previous research has reported elevated MIF levels in patients with schizophrenia (Liu et al. 2020). On the other hand, it is important to emphasize that the elevated MIF levels were detected in drug-naïve, first-episode patients and that 10 weeks of risperidone treatment led to a significant reduction in MIF concentration. If schizophrenia onset and *Toxoplasma* infection are indeed related, the reported elevation of MIF may reflect a transient response linked to the waning acute phase of infection rather than to latent infection. It is therefore plausible that in a cohort composed primarily of individuals with chronic infections lasting over a decade (originally diagnosed 10–13 years ago in a longitudinal study), the concentration of this molecule had already declined significantly.

The immunological hypothesis of schizophrenia posits that neuroinflammation plays a key role in the etiology of this severe mental disorder as well as in the expression of many of its symptoms (Aricioglu et al. 2016). It is virtually certain that the causes of neuroinflammation are diverse, and that latent toxoplasmosis may be only one of many contributing factors. Warm-blooded animals, including primates, serve as intermediate hosts for *Toxoplasma gondii*, facilitating the transmission of the parasite to its definitive host, felids, via predation (Tenter et al. 2000). From an evolutionary perspective, it is therefore in the parasite’s interest not to kill its host prematurely but rather to allow it to survive long enough to accumulate tissue cysts, ideally from genetically diverse parasite strains, thereby increasing the genetic diversity of gametes during sexual reproduction of the parasite in the feline host. Therefore, unlike other agents capable of inducing neuroinflammation (and thereby, potentially, schizophrenia), *Toxoplasma gondii* may have evolved specific adaptations to modulate neuroimmune processes in a way that preserves host viability. This may explain why the neuroinflammatory patterns observed in patients with schizophrenia – potentially arising from heterogeneous causes – differ in several respects from those associated with latent toxoplasmosis.

## Limitations

The primary limitation of this study was the relatively small sample size. However, the study followed a case–control design and was intentionally enriched for *Toxoplasma*-infected individuals to increase statistical power for detecting infection-related effects. Uninfected controls were selected from a larger cohort to match infected individuals in terms of age and sex, thereby minimizing potential confounding effects. While the sample size was sufficient to detect significant effects of infection on several biomolecules, other comparisons – despite showing substantial differences in mean concentrations between infected and uninfected individuals – did not reach statistical significance. This suggests that some biologically meaningful differences may have remained undetected due to limited statistical power. Future research, particularly if it follows an age-stratified design, would benefit from larger sample sizes.

The selection of biomarkers analyzed in this study was constrained by the composition of commercially available assay kits. As a result, some potentially relevant molecules could not be included. Furthermore, for certain biomarkers, the assay sensitivity was insufficient for reliable detection in a non-clinical population, leading to their exclusion from the statistical analyses.

A large proportion of the individuals willing to participate in our study had already taken part in other time-consuming, non-compensated studies more than a decade ago. The sample is therefore almost certainly not a random cross-section of the general population but is more likely composed of curious and altruistic individuals (as the rewards offered to participants were not substantial this time either). It is thus uncertain to what extent our findings can be generalized to the broader population. Nevertheless, given that we investigated concentrations of brain damage markers and cytokines, it seems unlikely that curiosity or altruism would systematically influence how these molecular parameters relate to *Toxoplasma* infection.

## Conclusion

This study tested whether previously reported differences in cytokine and brain injury biomarker levels between individuals with and without schizophrenia could partly reflect the higher prevalence of latent *Toxoplasma gondii* infection in the affected individuals. Our findings indicate that chronically infected individuals exhibit signs of mild but persistent neuroinflammation and neuro-immune dysregulation. Several observed changes – such as elevated S100B and KLK6 or trends toward increased FGF-21 – mirror those reported in schizophrenia. However, other findings differ markedly. For instance, we detected decreased MIF concentrations in infected subjects, while studies on schizophrenia have reported increased MIF levels, at least in early-stage, untreated patients. Taken together, our findings suggest that although latent toxoplasmosis may contribute to certain immune abnormalities seen in schizophrenia, it is insufficient to account for the full neuroinflammatory profile associated with the disorder. The observed immunological alterations in schizophrenia likely stem from a range of causes, and chronic toxoplasmosis represents only one possible contributing factor. Taken together, our findings do not support the hypothesis that the elevated prevalence of latent toxoplasmosis among patients with schizophrenia fully explains their neuroinflammatory profile, as the pattern observed in infected individuals diverges in several important respects.

## Data Availability

All data produced are available online at figshare 10.6084/m9.figshare.30408286

https://figshare.com/s/21941788d5c0591caf69

## Acknowledgment

The study was supported by the Czech Science Foundation supported this work (grant no. 22-20785S).

## Author Contributions statement

JF, ŠK and FŠ designed the study, JU, JT, MH, BŠ, JP, PN and JV performed the study, JF analyzed data, JF drafted the paper, and all coauthors participated on writing the paper.

